# Outcomes and Cardiovascular Comorbidities in a Predominantly African-American Population with COVID-19

**DOI:** 10.1101/2020.06.28.20141929

**Authors:** Ann B. Nguyen, Gaurav A. Upadhyay, Ben Chung, Bryan Smith, Stephanie A. Besser, Julie A. Johnson, John Blair, R. Parker Ward, Jeanne DeCara, Tamar Polonsky, Amit R. Patel, Jonathan Grinstein, Luise Holzhauser, Rohan Kalathiya, Atman P. Shah, Jonathan Paul, Sandeep Nathan, James Liao, Roberto M. Lang, Krysta Wolfe, Ayodeji Adegunsoye, David Wu, Bhakti Patel, Monica E. Peek, Doriane Miller, Dinesh J. Kurian, Stephen R. Estime, Allison Dalton, Avery Tung, Michael F. O’Connor, John P. Kress, Francis J. Alenghat, Roderick Tung

## Abstract

**Importance:** Racial disparities in COVID-19 outcomes have been amplified during this pandemic and reports on outcomes in African-American (AA) populations, known to have higher rates of cardiovascular (CV) comorbidities, remain limited.

**Objective:** To examine prevalence of comorbidities, rates of hospitalization and survival, and incidence of CV manifestations of COVID-19 in a predominantly AA population in south metropolitan Chicago.

**Design, Setting, Participants:** This was an observational cohort study of COVID-19 patients encountered from March 16 to April 16, 2020 at the University of Chicago. Deidentified data were obtained from an institutional data warehouse. Group comparisons and logistic regression modeling based on baseline demographics, clinical characteristics, laboratory and diagnostic testing was performed.

**Exposures:** COVID-19 was diagnosed by nasopharyngeal swab testing and clinical management was at the discretion of treating physicians.

**Main Outcomes and Measures:** Primary outcomes were hospitalization and in-hospital mortality, and secondary outcomes included incident CV manifestations of COVID-19 in the context of overall cardiology service utilization.

**Results:** During the 30 day study period, 1008 patients tested positive for COVID-19 and 689 had available encounter data. Of these, 596 (87%) were AA and 356 (52%) were hospitalized, of which 319 (90%) were AA. Age > 60 years, tobacco use, BMI >40 kg/m^2^, diabetes mellitus (DM), insulin use, hypertension, chronic kidney disease, coronary artery disease (CAD), and atrial fibrillation (AF) were more common in hospitalized patients. Age > 60 years, tobacco use, CAD, and AF were associated with greater risk of in-hospital mortality along with several elevated initial laboratory markers including troponin, NT-proBNP, blood urea nitrogen, and ferritin. Despite this, cardiac manifestations of COVID-19 were uncommon, coincident with a 69% decrease in cardiology service utilization. For hospitalized patients, median length of stay was 6.2 days (3.4-11.9 days) and mortality was 13%. AA patients were more commonly hospitalized, but without increased mortality.

**Conclusions and Relevance:** In this AA-predominant experience from south metropolitan Chicago, CV comorbidities and chronic diseases were highly prevalent and associated with increased hospitalization and mortality. Insulin-requiring DM and CKD emerged as novel predictors for hospitalization. Despite the highest rate of comorbidities reported to date, CV manifestations of COVID-19 and mortality were relatively low. The unexpectedly low rate of mortality merits further study.

**KEY POINTS:** *Questions:* What comorbidities are present in African Americans (AA) with COVID-19 and what are the associations with subsequent hospitalization and mortality? What is the incidence of COVID-19-associated cardiac manifestations requiring cardiology service utilization?

*Findings:* In this observational cohort study that included 689 patients with COVID-19 from south metropolitan Chicago (87% AA), cardiovascular (CV) comorbidities were highly prevalent and more common in those that required hospitalization. In addition to AA, age > 60 years, tobacco use, BMI >40 kg/m^2^, diabetes mellitus, hypertension, chronic kidney disease, coronary artery disease (CAD), and atrial fibrillation (AF) were more common in those hospitalized. Age > 60 years, tobacco use, CAD, and AF were associated with in-hospital mortality. Despite this, cardiac manifestations of COVID-19 were uncommon, and cardiology service utilization was low. In-hospital mortality was 13%. AA patients were more commonly hospitalized, but without increased mortality.

*Meaning:* In a predominantly AA population with COVID-19 at a major academic hospital located in south metropolitan Chicago, CV comorbidities were common and were risk factors for hospitalization and death. Although the highest rates of comorbidities to date were present in this cohort, mortality was relatively low and merits further study.

---

COVID-19 has swept across the globe at an unprecedented pace involving 185 countries, and has resulted in over 350,000 deaths worldwide and 100,000 deaths in the United States (US) so far.^1^ There is an urgent need to stratify risk for hospitalization, critical illness, and mortality on both medical and public health grounds during the current mitigation effort. Early reports of the pandemic from Italy and China do not reflect the demographic and disease burden of the US population.^2-5^ In the US, more than 40% of the population are obese,^6^ and racial diversity is the rule in large cities, which have high population density and high COVID-19 incidence. Disparities of health outcomes among African-Americans (AA) are well-documented for a range of acute and chronic diseases, and they are attributable to limited access to health care, socioeconomic factors, and long-standing structural inequalities.^7-11^ These health inequalities lay the foundation for the emergence of increased COVID-19 prevalence and mortality within the AA population, with states such as New York, Michigan, Louisiana, and Illinois reporting disproportionately higher deaths among AA than the general population.^7^

To date, however, there have been limited reports of COVID-19 outcomes from metropolitan centers with a predominantly AA population, known to have high prevalence of cardiovascular (CV) comorbidities. In two series reported from the New York City (NYC) area, AA represented the minority (<25%) of the cohort.^12,13^ There are only 2 published studies to date focusing on COVID-19 in populations where AA represent the majority of patients.^14,15^ Additionally, pre-existing CV disease has been proposed to compound risk during COVID-19 and isolated cases of new-onset cardiac manifestations such as myocarditis and ST segment elevation myocardial infarction (STEMI) have been reported.^16-21^ Despite multiple societal guidance statements discussing possible CV manifestations of COVID-19,^22-25^ the incidence of subsequent CV events is not well-defined.

We report the first experience from south metropolitan Chicago of patients admitted during the first 30 days of the COVID-19 pandemic to measure the rate of hospitalization and mortality amongst predominantly AA patients with COVID-19, examine the prevalence and spectrum of CV comorbidities and chronic diseases among these patients, and assess the incidence of new cardiac manifestations of COVID-19 in the context of overall cardiology service utilization.

## METHODS

### Study Design and Participants

This retrospective observational study was conducted at the University of Chicago Medical Center (UCMC), serving south metropolitan Chicago. The study was approved by the UCMC Institutional Review Board. Relative to metropolitan Chicago, the penetration of testing was high (**Figure 1**). Two-thirds of testing occurred in areas ≥ 90^th^ percentile for testing volumes in Illinois. On-site large platform testing for severe acute respiratory syndrome coronavirus 2 (SARS-CoV-2) via nasopharyngeal swab became available at UCMC on 3/16/20. All consecutive patients identified as positive between 3/16/20 (return of first positive test) to 4/16/20 were included. Final disposition of hospitalized patients was assessed through 5/25/2020.

**FIGURE 1.**
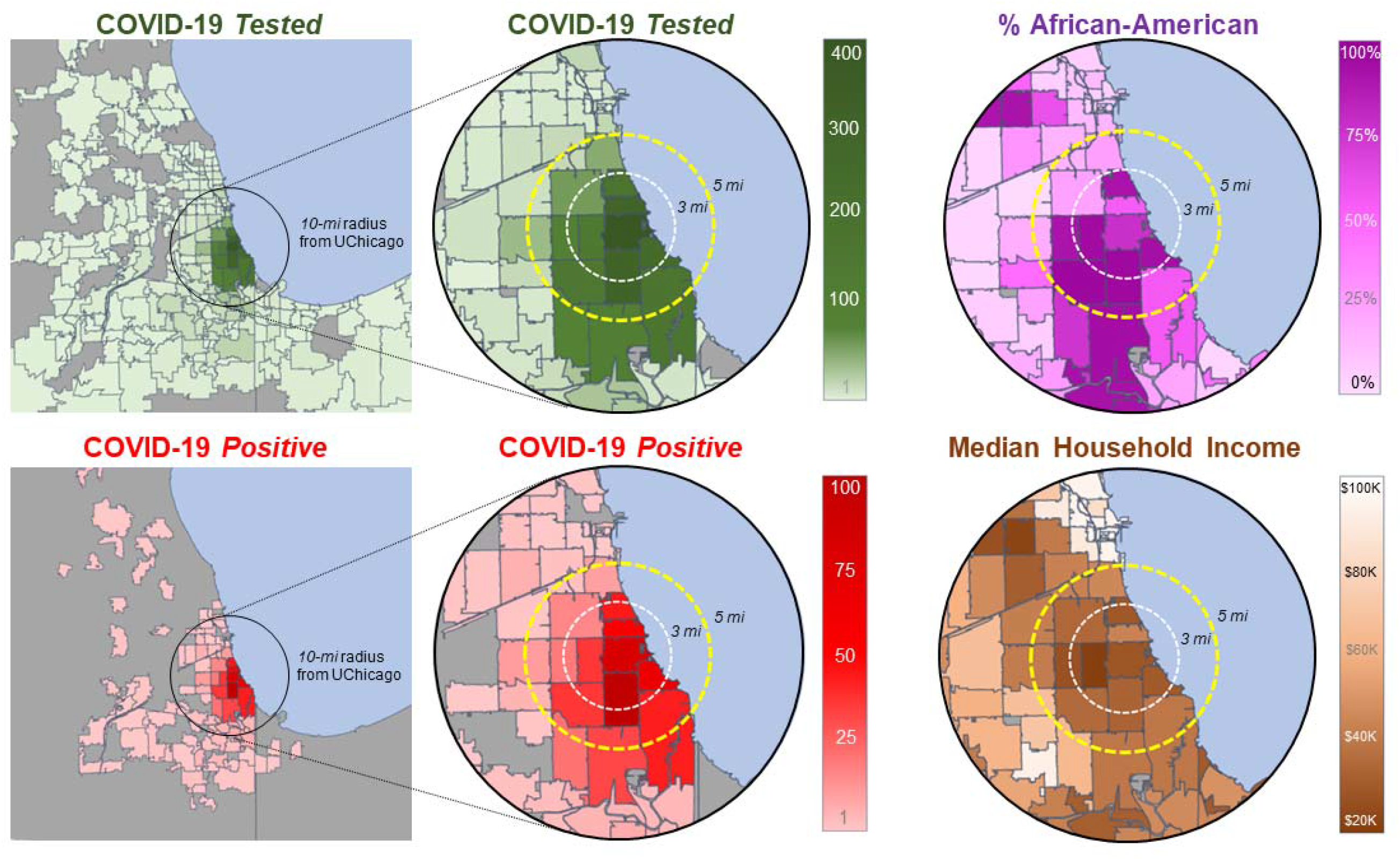
Geospatial maps of south metropolitan Chicago illustrating the density of COVID-19 testing, confirmed positive cases, proportion of African-Americans, and median annual income in the population studied.

### Data Collection

Data from this study were provided by the Clinical Research Data Warehouse (CRDW) maintained by the Center for Research Informatics (CRI) at the UCMC. The CRI is funded by the Biological Sciences Division and Institute for Translational Medicine/CTSA (NIH UL1 TR000430) at the University of Chicago. A limited, deidentified dataset of clinical information was acquired based on positive COVID-19 testing. The following data were recorded from patients treated at UCMC: baseline demographics, comorbidities based on ICD-10, outpatient medications, vital signs, laboratory values, complications, and treatment during hospitalization. Race data were collected by self-report in pre-specified categories, and comorbidities in the electronic medical record (EMR) were confirmed by history of previous provider encounters. Vital signs and laboratory values reflect the first available test results. For geospatial maps, zip codes were collected from COVID-19 patients, and race and income data of the community was curated from the US Census Bureau. In addition to those who died, the total number of COVID-19 patients that required hospitalization, admission to intensive care units (ICUs), and use of invasive ventilation were recorded.

### Outcomes

The primary endpoint was the rate of hospitalization and mortality with comparisons made between groups that did and did not meet these endpoints. Predictors of these outcomes were assessed by baseline demographics, comorbidities, and outpatient medications, and for those hospitalized, presenting vital signs and initial laboratory results.

The incidence of new CV manifestations directly related to COVID-19 was determined by new cases of left ventricular (LV) dysfunction or heart failure (HF), primary acute coronary syndrome, myocarditis, atrial fibrillation (AF), heart block, and ventricular arrhythmia (VA) requiring clinical intervention. Cardiology service utilization was assessed by the number of echocardiograms, cardiac catheterizations, electrophysiological procedures, and advanced HF census during the initial 30-day period.

### Statistical Analysis

Continuous baseline and clinical characteristics were expressed as medians with interquartile ranges and compared with Mann-Whitney U (Wilcoxon) tests after testing for normality using Shapiro-Wilk tests. Categorical baseline characteristics and medications were expressed as relative counts and percentages, and compared with Chi-square tests of association or Fisher exact tests. Denominators are provided in cases where data fields are incomplete from the EMR. Univariable logistic regression models were performed to determine univariable risk factors for hospitalization and in-hospital mortality. Baseline characteristics and outpatient medications with p ≤ 0.01 in the univariable regression models and present in at least 10 hospitalized patients were incorporated into a multivariable stepwise forward logistic regression model to calculate adjusted odds ratios (OR) and 95% confidence intervals for hospitalization. Independent parameters were checked for multicollinearity using Spearman rank correlations. Due to a low event rate, a multivariable logistic regression model was not able to be performed for in-hospital mortality. For cardiology service utilization, Wilcoxon rank sum matched-pair tests were used to assess differences in the 30-day study period compared to the previous 30 days for the following: incidence of echocardiographic studies, cardiac catheterizations, electrophysiological procedures, and HF census.

Tests were two-tailed and considered statistically significant with a p-value < 0.05. All statistical analyses were conducted using STATA MP version 15 (College Station, TX). Analysis began 4/17/20, and final mortality, ventilation status, and ICU status for hospitalized patients were collected through 5/25/20.

## RESULTS

During the first 30 days, 5,152 adults were tested at UCMC of whom 1,008 tested positive (**Supplemental Figure 1)**. Curbside tests were available to the general public at UCMC for a portion of the study period, and those curbside tests without any other concomitant data in the EMR were excluded from analysis (n=319). A total of 689 patients were analyzed and as of 5/25/20, 333 patients were treated and discharged from the emergency room or as outpatients, 356 patients were hospitalized, and 3 remained hospitalized.

Of the 689 patients, the median age was 55 years (40-68 years), 43% were male and 87% identified as AA (**Table 1**). Obesity (body mass index [BMI] > 30 kg/m^2^) was present in 57%, diabetes mellitus (DM) in 31%, current or past tobacco use in 26%, asthma in 16%, chronic kidney disease (CKD) in 12%, and chronic obstructive pulmonary disease (COPD) in 9%. Among CV comorbidities, history of hypertension (HTN) was present in 54%, coronary artery disease (CAD) in 15%, HF in 10%, and AF in 7%. Two or more CV comorbidities or risk factors were present in 56% of patients overall, in 81% with HTN, and in 81% with CKD. Outpatient medications included diuretics in 27%, statins in 22%, angiotensin-converting-enzyme inhibitor (ACEI) in 14%, angiotensin-receptor blocker (ARB) in 11%, beta-blockers in 21%, antiplatelets agents in 18%, and anticoagulation (vitamin K antagonists and direct-acting oral anticoagulants) in 13%. Compared to the minority of non-AA patients, AA patients were more likely to have severe obesity (BMI >40 kg/m^2^), HTN, DM, CAD, CKD, HF and COPD (**Supplemental Table 1**).

**TABLE 1.** Baseline patient characteristics and outpatient medications by hospitalization.

| Characteristic | All (n=689) | Hospitalized (n=356) | Not Hospitalized (n=333) | P |
| --- | --- | --- | --- | --- |
| Age median (IQR) | 55 (40-68) | 61 (50-73) | 47 (33-60) | <0.001 |
| Age Category |  |  |  | <0.001 |
| <40 | 171 (24.8) | 45 (12.6) | 126 (37.8) | <0.001 |
| 40-60 | 247 (35.9) | 122 (34.3) | 125 (37.5) | 0.37 |
| >60 | 271 (39.3) | 189 (53.1) | 82 (24.6) | <0.001 |
| Male | 296 (43.0) | 171 (48.0) | 125 (37.5) | 0.01 |
| Race |  |  |  | 0.01 |
| African-American | 596 (86.5) | 319 (89.6) | 277 (83.2) | 0.01 |
| White | 46 (6.7) | 23 (6.5) | 23 (6.9) | 0.82 |
| Other | 47 (6.8) | 14 (3.9) | 33 (9.9) | 0.002 |
| Tobacco Use | 171 (25.8) | 114 (32.8); n=348 | 57 (18.0); n=316 | <0.001 |
| Obesity |  |  |  | 0.01 |
| BMI <30 kg/m2 | 261 (43.5) | 139 (42.5); n=327 | 122 (44.7); n=273 | 0.59 |
| BMI 30-40 kg/m2 | 234 (39.0) | 117 (35.8); n=327 | 117 (42.9); n=273 | 0.08 |
| BMI >40 kg/m2 | 105 (17.5) | 71 (21.7); n=327 | 34 (12.4); n=273 | 0.003 |
| Diabetes Mellitus | 186 (30.6) | 127 (42.1); n=302 | 59 (19.3); n=306 | <0.001 |
| HTN | 331 (54.4) | 210 (69.5); n=302 | 121 (39.5); n=306 | <0.001 |
| Stroke/Cerebrovascular Disease | 36 (5.9) | 29 (9.6); n=302 | 7 (2.3); n=306 | <0.001 |
| CKD (Stage III-VI) | 72 (11.8) | 61 (20.2); n=302 | 11 (3.6); n=306 | <0.001 |
| COPD | 53 (8.7) | 35 (11.6); n=302 | 18 (5.9); n=306 | 0.01 |
| Asthma | 105 (17.3) | 46 (15.2); n=302 | 59 (19.3); n=306 | 0.19 |
| Cancer | 52 (8.6) | 37 (12.3); n=302 | 15 (4.9); n=306 | 0.001 |
| CAD | 90 (14.8) | 66 (21.9); n=302 | 24 (7.8); n=306 | <0.001 |
| HF | 62 (10.2) | 52 (17.2); n=302 | 10 (3.3); n=306 | <0.001 |
| AF | 44 (7.2) | 31 (10.3); n=302 | 13 (4.3); n=306 | 0.004 |
| Outpatient Medications |  |  |  |  |
| ACEI | 70 (13.5) | 58 (19.8); n=293 | 12 (5.3); n=226 | <0.001 |
| ARB | 57 (11.0) | 41 (14.0); n=293 | 16 (7.1); n=226 | 0.01 |
| Anticoagulation | 66 (12.7) | 52 (17.8); n=293 | 14 (6.2); n=226 | <0.001 |
| Immunomodulator | 85 (16.4) | 51 (17.4); n=293 | 34 (15.0); n=226 | 0.47 |
| Antiviral | 80 (15.4) | 39 (13.3); n=293 | 41 (18.1); n=226 | 0.13 |
| Beta-Blocker | 108 (20.8) | 80 (27.3); n=293 | 28 (12.4); n=226 | <0.001 |
| Calcium channel blocker | 103 (19.9) | 77 (26.3); n=293 | 26 (11.5); n=226 | <0.001 |
| Antiplatelet | 91 (17.5) | 69 (23.6); n=293 | 22 (9.7); n=226 | <0.001 |
| **NSAID** | 95 (18.3) | 47 (16.0); n=293 | 48 (21.2); n=226 | 0.13 |
| **Statin** | 115 (22.2) | 90 (30.7); n=293 | 25 (11.1); n=226 | **<0.001** |
| **Diuretics** | 138 (26.6) | 102 (34.8); n=293 | 36 (15.9); n=226 | **<0.001** |
| **Insulin** | 62 (12.0) | 52 (17.8); n=293 | 10 (4.4); n=226 | **<0.001** |
| **Asthma-COPD inhaler** | 155 (29.9) | 77 (26.3); n=293 | 78 (34.5); n=226 | **0.042** |

*Table Legend:* BMI – Body Mass Index; HTN – hypertension; CKD – Chronic Kidney Disease; COPD – Chronic Obstructive Pulmonary Disease; CAD – Coronary Artery Disease; HF – Heart Failure; AF – Atrial Fibrillation; ACEI – Angiotensin Converting Enzyme Inhibitor; ARB – Angiotensin Receptor Blocker; NSAID – Non-steroidal Anti-inflammatory Drugs

### Comorbidities in Hospitalized Patients

Of 689 COVID-19 patients analyzed, 52% required hospitalization. Hospitalized patients were more often >60 years and AA, with a higher prevalence of every comorbidity analyzed, except asthma (**Table 1**). Higher outpatient usage of ACEI or ARBs, beta-blockers, calcium channel blockers, statins, antiplatelet therapy, and anticoagulation was observed among hospitalized compared to non-hospitalized patients. Univariable testing of demographics and comorbidities showed highest ORs for CKD (6.79 [3.49-13.19]; p<0.001), HF (6.16 [3.07-13.37]; p<0.001), stroke (4.54 [1.96-10.53]; p<0.001), age >60 years (3.46 [2.50-4.79]; p<0.001), DM (3.04 [2.11-4.37]; p<0.001), HTN (3.49 [2.50-4.88];p<0.001) and CAD (3.29 [2.00-5.41]; p<0.001) for hospitalization (**Table 2**). AA patients had an OR of 1.74 (1.12-2.72; p=0.01) for hospitalization. Outpatient use of most major CV medications were significantly associated with hospitalization.

**TABLE 2.** Univariable and multivariable analysis for factors associated with hospitalization.

| Characteristic | Univariable Logistic Regression |  |  | Multivariable Logistic Regression (*n*=420) |  |  |
| --- | --- | --- | --- | --- | --- | --- |
|  | OR | 95% CI | p | OR | 95% CI | p |
| **CKD (Stage III-VI)** | 6.79 | (3.49-13.19) | <0.001 | 2.77 | (1.24-6.18) | 0.01 |
| **HF** | 6.16 | (3.07-13.37) | <0.001 |  |  |  |
| **Insulin** | 4.66 | (2.31-9.40) | <0.001 | 3.53 | (1.64-7.60) | 0.001 |
| **Stroke/Cerebrovasc Disease** | 4.54 | (1.96-10.53) | <0.001 |  |  |  |
| **ACEI** | 4.40 | (2.30-8.42) | <0.001 |  |  |  |
| **Statin** | 3.56 | (2.20-5.78) | <0.001 |  |  |  |
| **HTN** | 3.49 | (2.50-4.88) | <0.001 |  |  |  |
| **Age > 60** | 3.46 | (2.50-4.79) | <0.001 | 3.08 | (1.92-4.92) | <0.001 |
| **CAD** | 3.29 | (2.00-5.41) | <0.001 |  |  |  |
| **Anticoagulation** | 3.27 | (1.76-6.06) | <0.001 |  |  |  |
| **Diabetes Mellitus** | 3.04 | (2.11-4.37) | <0.001 |  |  |  |
| **Antiplatelet** | 2.86 | (1.70-4.79) | <0.001 |  |  |  |
| **Diuretics** | 2.82 | (1.83-4.33) | <0.001 | 1.75 | (1.06-2.90) | 0.03 |
| **Calcium Channel Blocker** | 2.74 | (1.69-4.45) | <0.001 |  |  |  |
| **Cancer** | 2.71 | (1.45-5.05) | <0.001 |  |  |  |
| **Beta-blocker** | 2.66 | (1.66-4.26) | <0.001 |  |  |  |
| **AF** | 2.58 | (1.32-5.03) | 0.01 |  |  |  |
| **Tobacco Use** | 2.21 | (1.54-3.19) | <0.001 |  |  |  |
| **ARB** | 2.14 | (1.17-3.91) | 0.01 |  |  |  |
| **BMI >40 kg/m<sup>2</sup>** | 1.95 | (1.25-3.04) | 0.003 | 2.84 | (1.58-5.13) | 0.001 |
| **African-American** | 1.74 | (1.12-2.72) | 0.01 |  |  |  |
| **Male** | 1.54 | (1.14-2.08) | 0.01 | 1.89 | (1.21-2.95) | 0.01 |
| **COPD/Asthma** | 1.02 | (0.70-1.48) | 0.93 |  |  |  |
| **Asthma/COPD inhaler** | 0.68 | (0.46-0.99) | 0.043 |  |  |  |

*Table Legend:* CKD – Chronic Kidney Disease; HF – Heart Failure; ACEI – Angiotensin Converting Enzyme Inhibitor; HTN – hypertension; CAD – Coronary Artery Disease; AF – Atrial Fibrillation; ARB – Angiotensin Receptor Blocker; BMI – Body Mass Index; COPD – Chronic Obstructive Pulmonary Disease

Multivariable analysis resulted in 6 baseline characteristics with significant OR for hospitalization: age >60 years (3.08 [1.92-4.92]; p<0.001), male sex (1.89 [1.21-2.95]; p=0.01), severe obesity (BMI >40 kg/m^2^) (2.84 [1.58-5.13]; p=0.001), CKD (2.77 [1.24-6.18]; p=0. 01), outpatient insulin (3.53 [1.64-7.60]; p=0.001) and diuretic use (1.75 [1.06-2.90]; p=0.03) (**Table 2**).

### In-Hospital Mortality

Of patients who were hospitalized, 44% required critical care during hospitalization, and 13% died. Among patients receiving critical care, 45% required invasive ventilation and 27% died. Mortality in invasively ventilated patients was 46%. Patient characteristics, vital signs, and laboratory data by mortality status are shown in **Table 3**.

**TABLE 3.**
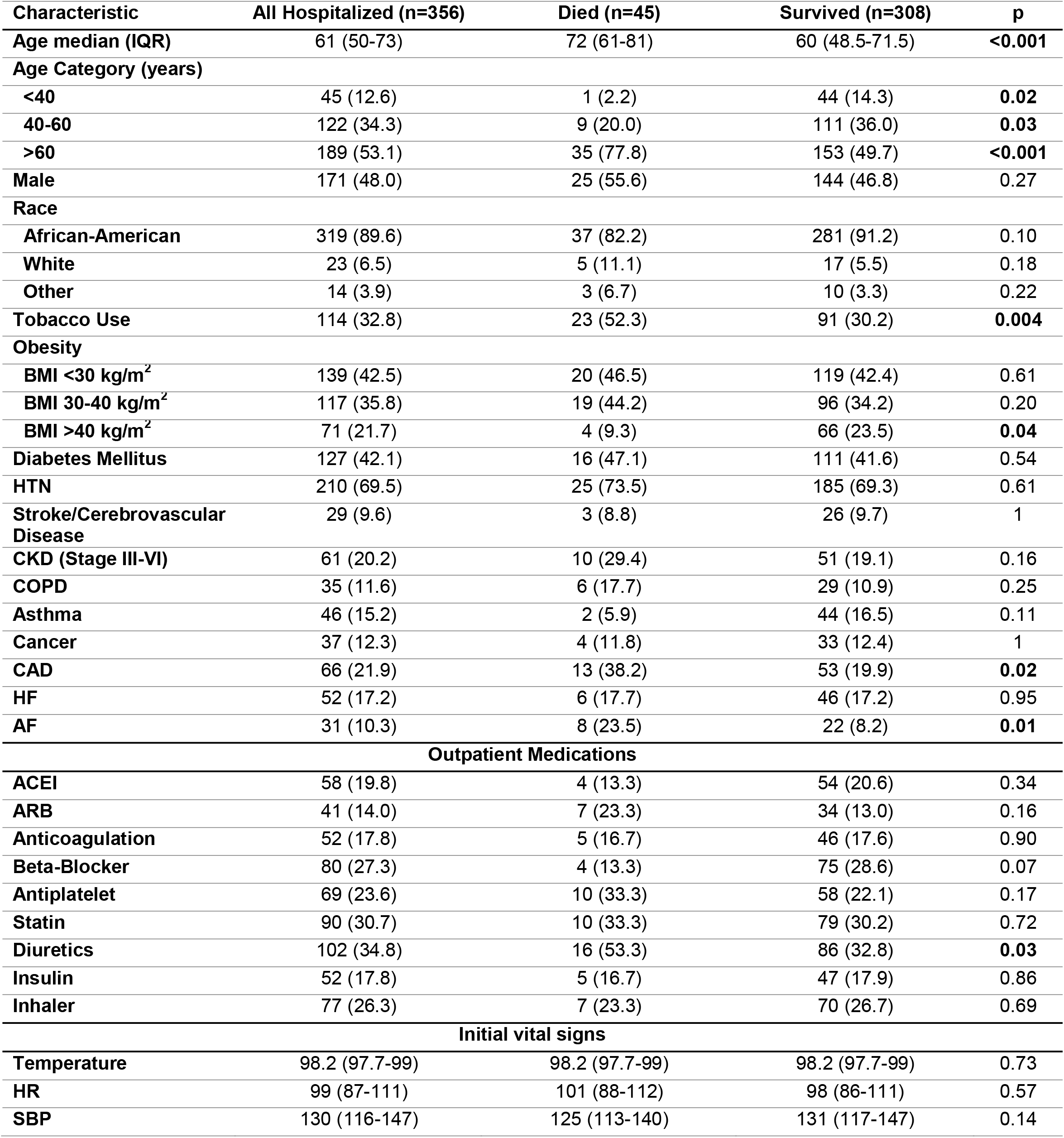
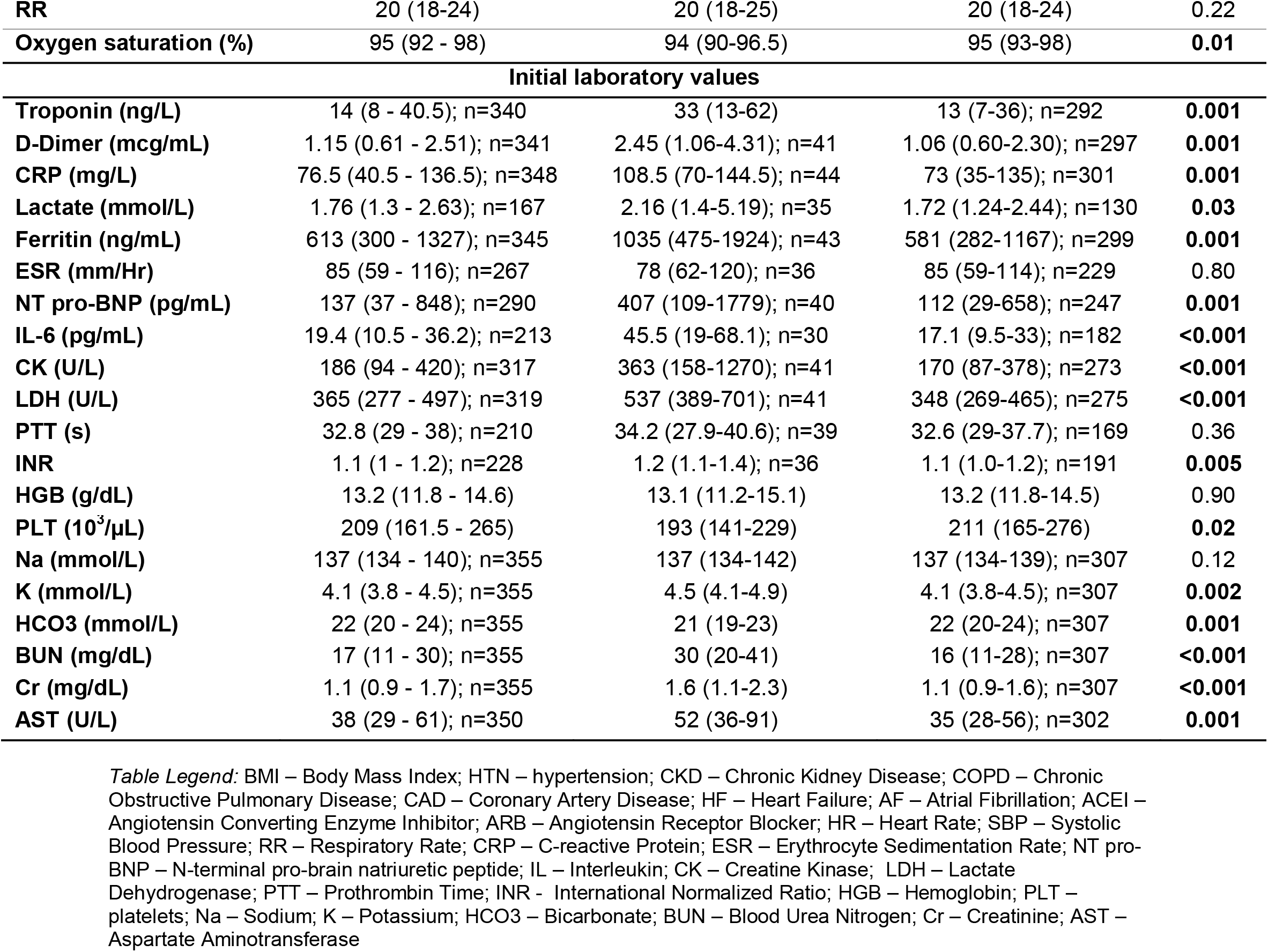
Patient characteristics, outpatient medications, initial vital signs, laboratory values by survival.

Patients who died were more often >60 years compared to those who survived (78% vs 50%, p<0.001), and only 1 patient less than 40 years died (**Table 3**). There was no difference in the prevalence of AA patients between those who died and survived (82% vs 91%; p=0.10) Patients who died were more likely to have history of tobacco use (52% vs 30%, p =0.004), CAD (38% vs 20%, p=0.02) and AF (24% vs 8%, p=0.01). Although severe obesity (BMI >40 kg/m^2^) was a multivariable predictor of hospitalization, it was not associated with increased in-hospital mortality (9% vs 24%, p=0.04).

Among initial vital signs and laboratory data, lower oxygen saturation and higher levels of creatinine, D-dimer, C-reactive protein, lactate, aspartate aminotransferase, ferritin, interleukin-6, and lactic dehydrogenase were more frequently observed in those who died compared to those who survived (all p < 0.05).

Elevated initial values of cardiac biomarker NT-pro-BNP were observed in those who died (median 407 [109-1779] vs 112 [29-658] pg/mL, p=0.001). Elevated initial high-sensitivity troponin (>22 ng/dL) values were observed in 41% of hospitalized, 57% of patients requiring critical care, and 67% of those who died. Higher elevations in median initial troponin were also observed in those who eventually died (33 [13-62] vs 13 [7-36] ng/L; p=0.001).

Univariable testing of demographics, comorbidities, initial vital signs and initial laboratory values showed high ORs for age >60 years (3.55 [1.70-7.41]; p<0.001), tobacco use (2.53 [1.33-4.80]; p=0.005), CAD (2.50 [1.18-5.31]; p=0.02), AF (3.43 [1.39-8.47]; p=0.008), troponin >22 ng/L (3.36 [1.73-6.52]; p<0.001), BUN >20 mg/dL (4.72 [2.34-9.51]; p<0.001), and NT-proBNP > 125 pg/mL (3.07 [1.54-6.12]; p=0.001) for death (**Supplemental Table 2**). BMI >40 kg/m^2^ was not associated with mortality (0.33 [0.12-0.97]; p=0.044). AA (0.44 [0.19-1.05]; p=0.07) and initial vital signs were not significantly associated with mortality.

### Cardiac Manifestations of COVID-19 and Cardiology Service Utilization

From the pre-COVID-19 (2/17-3/15/20) to the COVID-19 period (3/16-4/16/20), cardiology service utilization decreased significantly (**Figure 2**). The number of echocardiograms declined by 69% (80 [78-84] to 25 [21-27] per day, p<0.001) due to efforts to minimize exposure of equipment and personnel. Echocardiograms were performed on the basis of clinical necessity and approved by the director. One echocardiogram showed known left ventricular (LV) dysfunction with ventricular assist device; another showed a clinically significant pericardial effusion. There were no observed cases of new-onset LV dysfunction (ejection fraction < 50%) observed during this time period among those with echocardiography performed. Interventional catheterization procedures decreased by 65% (9 [7-10] to 3 [2-3] per day, p<0.001). Electrophysiology procedures decreased by 78% (5 [4-5] to 1 [0-2] per day, p<0.001). A total of three catheterization procedures were performed on COVID-19 patients: one patient with shock underwent a left and right heart catheterization and was found to have non-obstructive coronary disease; one patient with non-STEMI underwent angiography that showed non-obstructive coronary disease; and one heart transplantation patient required pericardiocentesis for pericardial effusion. There were two cardiology consults for syncope and supraventricular tachycardia. No cases of STEMI, VA, or heart block requiring procedural intervention were observed related to COVID-19. Right heart catheterizations for HF decreased by 40% (5 [4-7] to 3 [2-4] per day, p<0.001). The daily census of advanced HF patients decreased by 67% (18 [16-18] to 6 [4-7], p<0.001). There were no consultations for advanced HF therapies or mechanical circulatory support related to COVID-19.

**FIGURE 2.**
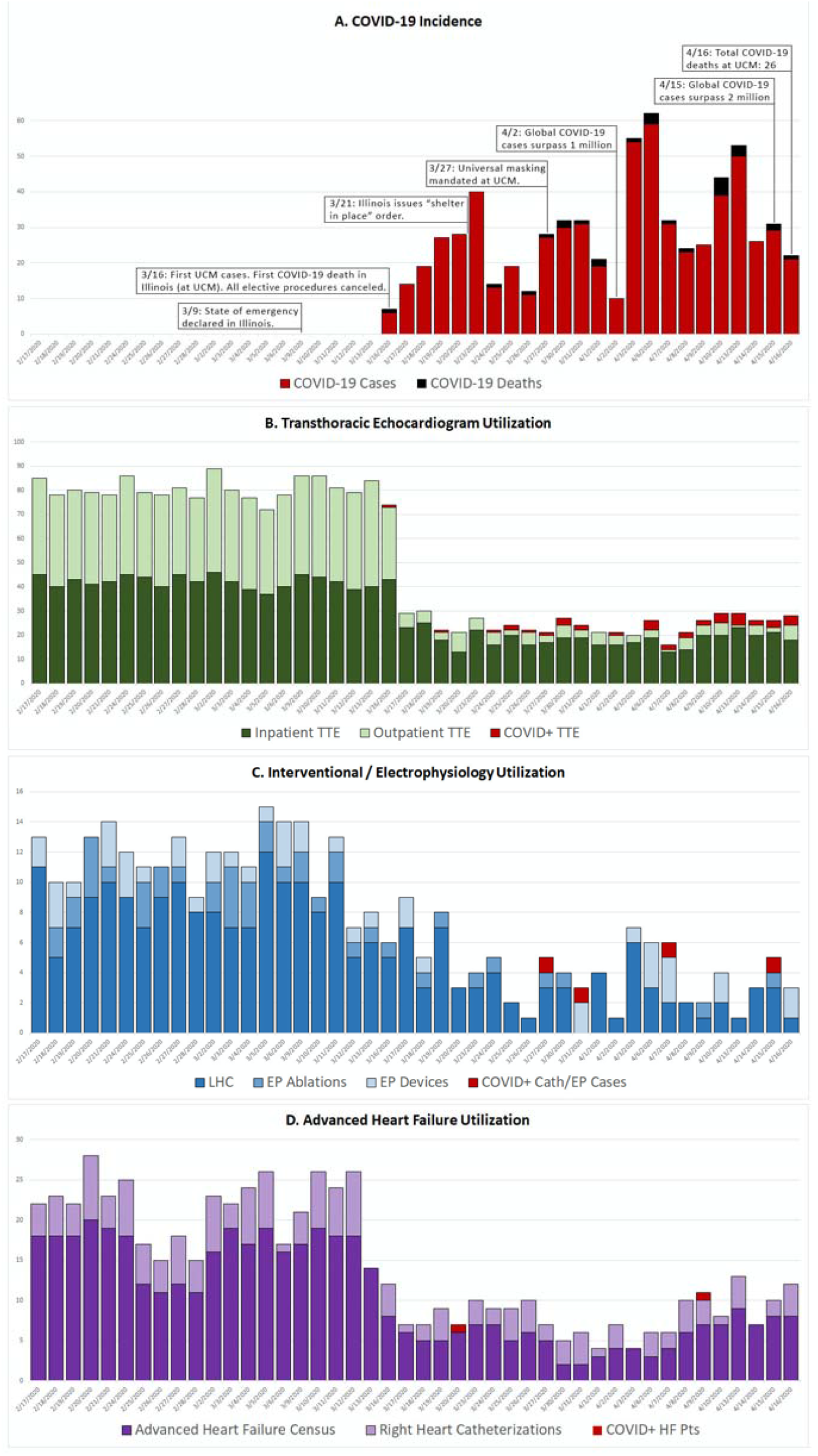
Case counts of confirmed COVID-19 cases over the study period compared to the prior 30 days with cardiology service line utilization by procedural volume, echocardiographic studies, and heart failure inpatient census. Testing on a lower number of confirmed COVID-19 cases are indicated in red.

## DISCUSSION

This investigation has the highest majority of AA patients with COVID-19 to date. Health care disparities have been accentuated during the COVID-19 pandemic and the root causes are multifactorial.^26-29^ Though AA currently make up 30% of the population of Chicago, they comprise ∼50% of COVID-19 cases and ∼70% of COVID-19 deaths.^27,30^ This experience from south metropolitan Chicago is unique in that AA comprised 90% of hospitalized patients and testing was performed in areas ≥ 90^th^ percentile for testing volumes in Illinois. As AA are disadvantaged by limited access to healthcare, the present data explore outcomes from a community that received earlier testing and access to advanced therapies at an academic hospital specializing in tertiary care. In addition to AA and age > 60 years, CV comorbidities and CV disease equivalents such as obesity, tobacco use, DM, HTN, stroke, CKD, CAD, HF, and AF were more prevalent in those requiring hospitalization. Furthermore, smoking history, CAD, and AF were more common in those who died and associated with in-hospital mortality on univariable analysis. Consistent with multiple reports, the prevalence of CV comorbidities and chronic disease was disproportionately high in AA patients.^10,31^

Relative to other early populations reported from China, Italy, and NYC,^2-5,12,13,32^ our hospitalized COVID-19 population had the highest rate of CV comorbidities and other chronic medical illnesses to date. This remains true even when compared to more recently published reports of predominantly AA COVID-19 patients from Georgia and Louisiana.^14,15^ Over 50% of patients suffered from obesity and HTN, 42% had DM, 22% had CAD, 12% had COPD, and 15% had asthma (**Supplemental Table 3**). Obesity, HTN, CAD, and DM are chronic disease states with a disproportionately high prevalence in the AA community and serve as major pathogenic mechanisms for accelerated development of CKD. CKD is a well-recognized risk factor for morbidity and early death, and has been uncommon in larger scale COVID-19 cross-sectional reports of non-nursing home patients (1-10%), and was present in 20% of our cohort of hospitalized patients.^12,13,33^

While DM has been reported as a risk factor for adverse outcomes with COVID-19, we found that outpatient insulin therapy was associated with the highest risk for hospitalization on multivariable testing, which has not been previously reported. Patients with insulin-requiring diabetes have more poorly controlled glycemic states and are at higher risk for ketoacidosis and hyperosmolar states in the setting of infection.

Outpatient diuretic use, another independent risk factor, is used both in the treatment of HTN and HF. With regard to outpatient medications, ACEI and ARB usage did not appear to portend improved prognosis as suggested by some reports from China, although overall use remained limited in the study population.^18,34,35^ Of all CV comorbidities, CAD and AF were more common in those who died and were associated with in-hospital mortality on univariable analysis. AF is well known as a harbinger of multiple disease processes and could represent those with highest disease burden and thus risk for death. Finally, though AA patients were more commonly hospitalized, they did not have higher rates of in-hospital death, consistent with previous reports.^14,15^

CV manifestations of COVID-19 have been reported in aggregated case series,^16,17,19-21^ and multiple societal guidance statements^22,23^ have alerted the community of the possibility of myocarditis, STEMI, dysrhythmia, and heart block. In this investigation, we found a low incidence of new CV manifestations related to COVID-19 by screening overall cardiology service line utilization. System-wide changes in practice reduced the incidence of consultation and routine testing, but the overall case volume and clinical demand across all cardiac sub-specialties in response to COVID-19 remained low. With with low rates of need for cardiology services, our findings suggest that cardiac involvement related specifically to SARS-CoV-2 may currently be overrepresented in the literature, and fulminant myocardial injury with viral involvement appears uncommon.^19^ With that noted, even relatively modest elevations of troponin were associated with mortality in the present study, and may be more valuable than biomarkers such as CRP and D-dimer which were uniformly elevated regardless of eventual outcome.

### LIMITATIONS

This report is limited by the single-center nature and small sample within the first 30 days of the first confirmed case of COVID-19. Although UCMC primarily functions to serve the local community during the COVID-19 pandemic, the generalizability may be limited by the tertiary nature of care along with the ability to rapidly implement hospital-wide protocols (i.e. negative pressure ventilation single occupancy, modified thromboembolic prophylaxis). Criteria and thresholds for hospitalization, admission to ICU, and invasive ventilation were dynamic over this time period of rapid adaptation to the crisis.

Many patients (∼40%) hospitalized received experimental pharmacologic therapy (hydroxychloroquine, remdesivir, lopinavir/ritonavir, tocilizumab) and the outcomes from these individualized and compassionate-use interventions are the subject of ongoing investigation. Presently, there is no conclusive evidence to support the efficacy of these therapeutic agents.^36-42^ Additionally, adjunctive and evolving strategies such as prone^43^ and helmet ventilation^44^, higher dose prophylactic anticoagulation, high-flow supplemental oxygen to delay invasive ventilation may have impacted the observed mortality rates and merit further investigation. Studies beyond the scope of this descriptive report are warranted to explain the lower observed mortality in this population with high proportion of comorbidities (**Supplemental Table 3**). While the impact of early citywide lockdown and access to large platform testing is difficult to measure, newly confirmed cases at UCMC appeared at a slower rate than that reported in NYC, and likely contributed to lower mortality rates. Additionally, at our institution, there was no shortage of ventilators during the study period and ICU capacity was adequate throughout, and in addition, definitions of critical care may vary between institutions. Finally, the incidence of subclinical myocarditis may be underreported as uniform performance and reporting of point-of-care ultrasound and laboratory testing were not mandated.

## CONCLUSIONS

In this AA predominant experience from south metropolitan Chicago, CV comorbidities and chronic disease states were highly prevalent and associated with hospitalization and mortality. In addition to previously reported risk factors (male sex, advanced age), insulin-requiring DM, obesity, and CKD emerged as novel predictors of hospitalization. Patients who died had higher rates of smoking history, CAD, and AF. Despite the highest rate of co-morbidities reported to date, CV manifestations of COVID-19 and mortality were relatively low. The unexpectedly low rate of mortality merits further study.

## Data Availability

Deidentified data will be made available upon request to corresponding authors and subject to approved institutional data use agreement.

## ACKNOWLEDGEMENTS

We would like to thank William Parker, MD, MS for review of this manuscript.

## SOURCES OF FUNDING

There are no sources of funding relevant to this work.

## DISCLOSURES

All authors report no conflicts.

## Notes

### Competing Interest Statement

The authors have declared no competing interest.

### Funding Statement

There was no funding for the work presented.

### Author Declarations

The University of Chicago IRB approved this study.

